# Factors affecting first return to work following arthroscopic rotator cuff repair: A retrospective observational study in an occupational medicine clinic and single-surgeon practice

**DOI:** 10.1101/2021.09.20.21263877

**Authors:** Ijaz Khan, Manaal Fatima, Corey Scholes, Vikram Kandhari, K.M. Ponnanna, Jonathan Herald

## Abstract

**Aims:** Determine factors affecting first return to work (RTW) status and time in patients treated with arthroscopic rotator cuff repair (aRCR) under state-based compensation schemes in New South Wales, Australia, compared to those outside such schemes.

**Material and Methods:** Patients undergoing aRCR by one surgeon with minimum 1-year follow-up were grouped into those under (CP) or outside (non-CP) workers or vehicle accident compensation schemes, matched by age and gender. RTW status and time were assessed using chi-square analysis and multivariable linear regression.

**Results:** Of 1054 available patients, 90 CP patients were identified with 29 consented and matched to non-CP (N=29). A higher proportion of CP patients (17.2 vs 0%, P<0.001) never returned to work and a lower proportion resumed pre-injury duties at first RTW (3 vs 52%, P<0.01). Median time to first RTW did not differ between CP and non-CP groups (5.1 vs 4.4, P=0.86). Smoking (P=0.007) and post-injury activity level (P=0.004) were significantly associated with longer time to first RTW, whereas compensation status was not.

**Conclusions:** CP patients undergoing aRCR in NSW are at risk of not returning to work. For those that do return, there is no significant difference compared to non-CP in time to first RTW. Particularly, patient and management factors associated with extended time to first RTW have been identified. Interventions aimed at modifiable factors such as smoking cessation and increasing preoperative activity may improve future outcomes.

## Introduction

Work-related injury and disease cost the national economy $AUD61.8billion annually [1], with shoulder conditions from body stress accounting for >$AUD4.2billion [2]. The proportion of claims due to symptomatic rotator cuff (RC) tears is up to 20% [3], and arthroscopic repair (aRCR) is a primary treatment option [4]. While treatment outcomes with respect to tendon healing and function are extensively reported [5,6], there is a dearth of information regarding modifiable factors for optimising return to work (RTW) after aRCR due to workplace injury.

RTW time and status after aRCR for work-related injuries are governed by patient and contextual factors. Compensable patients have reported significantly worse outcomes up to one year postoperatively [7]. Low scores on the Disabilities of the Arm, Shoulder and Hand (DASH) persist when controlled for age, sex, comorbidities, smoking, marital status, education, symptom duration, work demands, expectations and tear size. Six-month American Shoulder and Elbow Surgeons (ASES) and Western Ontario Rotator Cuff (WORC) Index scores were lower in compensable patients, even when adjusted for differences in age, gender, smoking status, baseline scores, symptom duration, injury type and associated biceps disorder [8].

The relationship between postoperative outcomes and RTW time, however, remains unclear. In one study, 89% of compensable patients returned to preoperative work 7.6 months after aRCR, and reported good outcomes on validated scoring scales but inferior subjective outcomes compared to uncompensated patients [9]. Older age, private sector employees, and part-open or arthroscopy cases prevented RTW in any capacity, while the number of injured tendons increased time away from full-time work but did not prevent RTW [10]. Female gender, heavy manual labour and postoperative bursitis may also prevent RTW [11].

In a Belgian study (N=73), highly compensated patients took significantly longer time off work compared to those receiving lower compensation (7 vs 2.5 months) [12]. A significant relationship was found between compensation level and physical work demand, with highest compensated patients holding jobs with higher physical demands. Facilitating RTW in the broader context of work-related injury and occupational disease requires a multidisciplinary approach, with age, gender, injury type, intervention duration, employer interest and employee motivation found to affect RTW (N=9850) [13]. Participants with interested employers were 23 times more likely to RTW than those without, and those with longer intervention periods (>5 months) were less likely to RTW.

From the perspective of RTW stakeholders (employers, insurers, lawyers and healthcare providers) in Australia, the factors rated as having the greatest influence on RTW were predominantly psychosocial and modifiable [14]. These included self-efficacy, postoperative psychological status, employer support and capacity to modify roles, recovery expectations, mood disorders and postoperative pain level. Establishing predictors for prolonged RTW from Australian compensation claims has been unsuccessful, given the design of claim forms and poor data quality [15].

While compensable patients typically report poorer outcomes and take longer to RTW than their non-compensated counterparts, there is considerable variation in the definitions of RTW status, which may be reported as first, final, pre-injury duties, part-time, same role, same employer but modified role, or otherwise unspecified. This presents a challenge when comparing RTW as an outcome between studies. Furthermore, whether this holds true in the Australian population remains unexplored, and the effect of modifiable factors on first RTW status and time away from work in this population are unclear. This study aims to determine the patient, pathology, treatment and postoperative management factors associated with RTW status and RTW time in patients presenting with RC pathology treated with aRCR under an Australian workers compensation scheme, compared to patients treated outside such schemes.

## Methods

### Patient selection and group matching

Patients who presented to an occupational health clinic diagnosed with an RC tear and elected to undergo aRCR by a single fellowship-trained surgeon with minimum one-year follow-up were retrospectively analysed. Patients indemnified under the New South Wales (NSW), Australia workers or motor vehicle accident compensable schemes (compensable patients; CP), were age- and gender-matched to controls treated outside any compensable scheme (non-compensable patients; non-CP). Exclusion criteria included patients withholding consent, were not contactable, had associated glenohumeral arthritis or shoulder instability with RC tear, or had previous pathology in the same shoulder (managed operatively or non-operatively). Non-CP patients were additionally excluded if they were non-privately funded. The non-CP group was matched to the CP group by age (within 5% range at first pass, and 10% range at second pass) and gender. Ethical clearance for this study was obtained from Bellberry Limited (HREC 2018-07-597-A-1) and the study was conducted in accordance with the National Statement for Ethical Conduct in Research (AUS) and the Declaration of Helsinki (2013 Rev).

### Surgical procedure

All patients received an aRCR in lateral position using standard posterior, lateral and anterior portals. Majority of patients underwent single row repair and in patients where the cuff was mobile and could be pulled over the lateral side of foot print, double row repair was performed. Similar perioperative anaesthesia and pain management protocols were followed for all patients, and all had subacromial decompression, acromioplasty and biceps tenodesis. Distal clavicle excision was not performed. Similar postoperative pain management and rehabilitation protocols were followed (see Supplementary Text), and all patients received follow-up from the same occupational health clinic with monitored physiotherapy during follow-up.

### Recruitment and data collection

Patients who met criteria were identified from the occupational health clinic and surgeon’s database and clinical records and then contacted to provide consent. Two controls per compensable patient were matched to account for withdrawal of consent or lack of contact. A phone follow-up was performed to collect data on: patient factors (marital status, education level, surgery side); contextual factors (employer at injury time, previous unemployment periods) and postoperative activity level (Tegner score). The Tegner Activity Scale [16] is a graduated activity list of daily living, recreation and competitive sports. Patients select a level of participation that best describes their current activity level and are assigned scores from 0 (worst) to 10 (best). Though it was developed for assessment of ACL injuries, the scale has been used to assess activity improvements in patients with RC injuries [17,18].

Electronic data was extracted from medical databases of the occupational health clinic and orthopaedic surgeon, and included: patient factors (age, gender, weight, height, postcode, injury side, smoking status, comorbidity, medications, substance abuse, alcohol consumption and mental health status); contextual factors (occupation, pre-injury employment status, physical work demand, previous claims, and durations between symptoms, presentation, claim and surgery); pathological factors (diagnosis, injury cause, previous treatments, tear characteristics, and baseline function); operative factors (date and location of surgery, repair type, surgical approach, surgery adjuncts, anaesthesia, analgesia, postoperative bursitis and rehabilitation protocol); and outcomes (RTW status and RTW time). The phone follow-up was performed by a fellow to avoid bias, and sensitive data such as substance abuse and mental health status were recorded as binary (yes or no) in the interest of privacy and to encourage compliance.

### Statistical analysis

A STROBE diagram [19] indicated the analysis workflow, with reasons for data exclusion, group identification and basic descriptives identified. Data was prepared for statistical analysis by examining completeness and outliers. For outliers, source material was re-examined to rectify transcription errors. Continuous variables were assessed for normality using Anderson-Darling tests and summarised by median and interquartile range (IQR). List-wise deletion was used for missing values in univariate analyses. Matched pairs were compared for continuous variables using one-sample Wilcoxon t-test and for 2×2 comparisons of categorical data using McNemar’s test of association. Categorical variables with multiple responses were compared between groups using chi-squared analysis. Box-Cox transformation with optimal lambda was used to restore surgery to RTW duration to a normal distribution and a forwards-backwards stepwise regression model was applied to associate patient characteristics, injury and treatment factors with time to RTW. Alpha was set at 0.15 for predictor inclusion into the model and for removal. Adjusted R^2^ was used to assess model fit and alpha of 0.05 was deemed critical for all tests. All statistical analyses were performed in a specialised software package (Minitab v18, PA, USA).

## Results

### Patient characteristics

Patients identified from the occupational health clinic and surgeon’s database as available for assessment (N=1054) were divided into CP and non-CP groups. Two groups of 29 patients undergoing aRCR between October 2007 and May 2018 were age- and sex-matched and included in the final analysis (Figure 1).

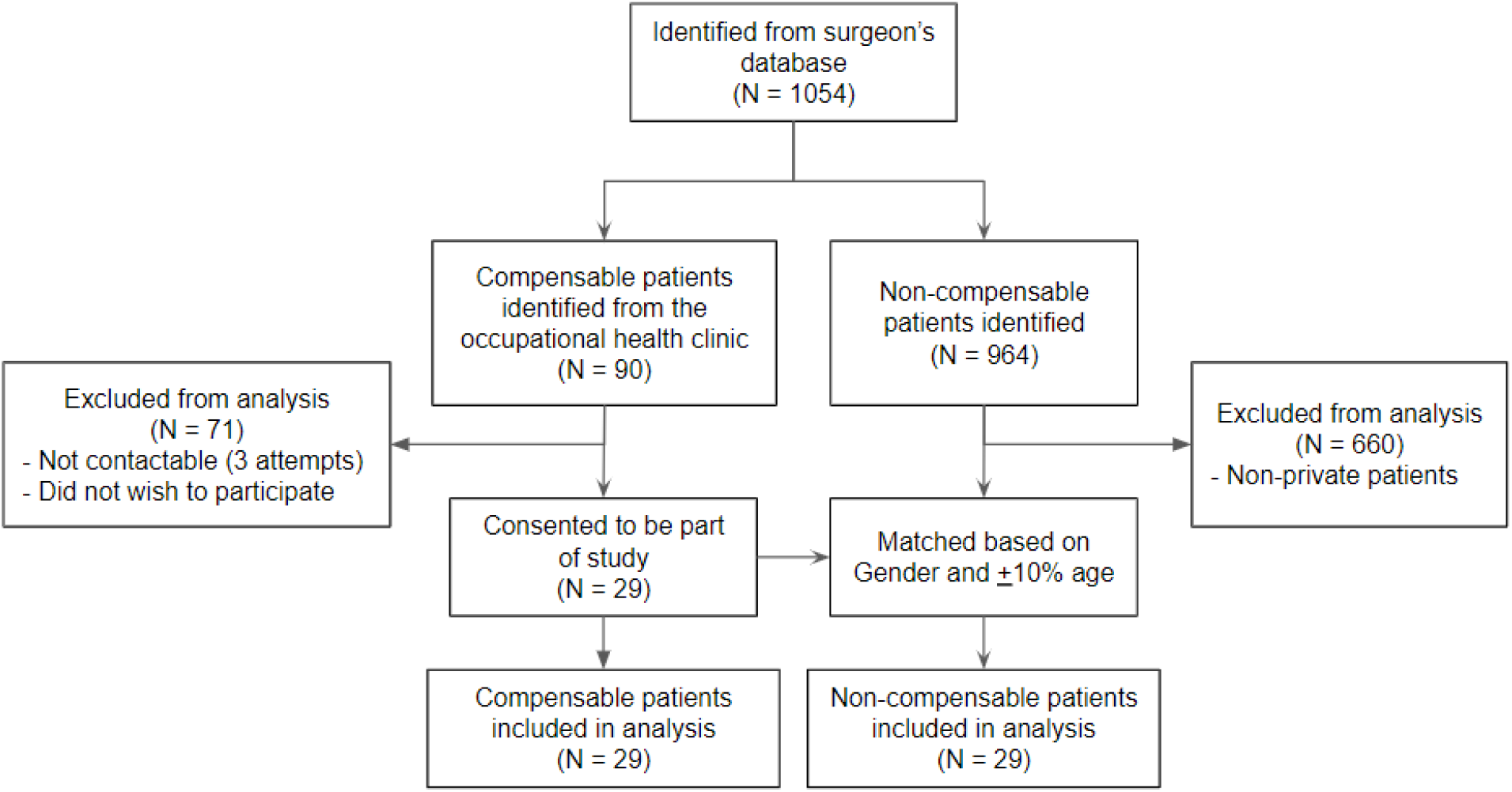

Median age at surgery was 54 (IQR 51-60) years for the CP and 59 (IQR 51-64) for the non-CP group, with 86% of the sample male. Bilateral injury was present in 14% of patients and 29% presented with comorbidities, with hypertension being the most frequently reported (35%). Patient characteristics differed significantly between the groups for age at surgery, occupation, pre-injury employment, education level and pre-injury activity (Table 1). No patients had any recorded mental health issues or substance abuse. The non-CP group made no previous compensation claims for other injuries, while 86% of the CP group had made up to five other claims. The CP group were compensated under the NSW workers compensation scheme, with only one patient compensated under third party insurance.

**Table 1:** Patient characteristics separated by compensation status

| Patient characteristics | Missing data | CP (N=29) | Non-CP (N=29) | All (N=58) | P-value |
| --- | --- | --- | --- | --- | --- |
| Age at surgery (median, IQR) | 0 | 53.5 (50.6-60.3) | 59.3 (51-63.6) | 55 (51-61.8) | <0.001 |
| Bilateral (%) | 0 | 20.7 | 6.9 | 13.8 | 0.12 |
| Comorbidities (%) | 0 | 31 | 27.6 | 29.3 | 0.77 |
| Currently smoke (%) | 0 | 17.2 | 10.3 | 13.8 | 0.44 |
| **Alcohol consumption (%)**<br>≥ 2 standard drinks per day | 0 | 0 | 10.3 | 5.2 | - |
| **Occupation (%)**<br>Heavy manual<br>Light manual<br>Office<br>Domestic duties | 0 | 65.5<br>27.6<br>6.9<br>0 | 10.3<br>27.6<br>41.4*<br>20.7 | 37.9<br>27.6<br>24.1<br>10.4 | **<0.001** |
| **Pre-injury employment status (%)**<br>Full-time<br>Part-time<br>Casual<br>Unemployed | 0 | 75.9<br>13.8<br>10.3<br>0 | 65.5<br>17.2<br>0<br>17.2* | 70.7<br>15.5<br>5.2<br>8.6 | **0.01** |
| **Currently married (%)** | 0 | 86.2 | 89.7 | 87.9 | 1 |
| **Education level (%)**<br>Post graduate<br>University degree<br>TAFE certificate/Diploma<br>Year 12<br>Year 10 or below | 0 | 0<br>13.8<br>3.5<br>62<br>20.7* | 10.3<br>31.1<br>10.3<br>44.8<br>3.5 | 5.2<br>22.4<br>6.9<br>53.4<br>12.1 | **0.02** |
| **Hand dominance on surgery side (%)** | 0 | 44.8 | 69 | 56.9 | 0.06 |
| **Pre-injury Tegner (median, IQR)** | 0 | 5 (4.5-5) | 5 (5-6) | 5 (5-6) | 0.092 |
| **Post-injury pre-surgery Tegner (median, IQR)** | 0 | 4 (3-5) | 5 (5-6) | 5 (3.3-5) | **<0.001** |

\*largest contribution to $\chi^2$ ; CP = compensable patients; non-CP = non-compensable patients

### Injury and treatment characteristics

No patients had involvement of the Teres Minor tendon, however the non-CP group presented with a significantly higher proportion of full-thickness tears compared to the CP group, and more were on leave at the time of diagnosis (Table 2). The CP group displayed significantly higher rates of work as a cause of injury, partial tears, incidence of concomitant shoulder pathology and longer period from first presentation to surgery (Table 2).

**Table 2:** Injury and treatment characteristics of RC repair cases separated by compensation status

| Injury and treatment characteristics | Missing data | CP (N=29) | Non-CP (N=29) | All (N=58) | P-value |
| --- | --- | --- | --- | --- | --- |
| **Goutallier Grade (%)**<br>1<br>2<br>3 | 0 | 89.7<br>6.9<br>3.4 | 86.2<br>6.9<br>6.9 | 87.9<br>6.9<br>5.2 | 0.836 |
| **Structures involved (%)**<br>Supraspinatus tendon<br>Infraspinatus tendon<br>Subscapularis tendon<br>Biceps | 0 | 96.5<br>13.8<br>31<br>55.2 | 100<br>27.6<br>31<br>55.2 | 98.3<br>20.7<br>31<br>55.2 | 1<br>0.39<br>1<br>1 |
| **Partial Tear (%)** | 0 | 37.9 | 13.8 | 25.9 | **0.039** |
| **Tear size (mm) (median, IQR)** | 0 | 12 (6-15.5) | 14 (10-24.5) | 12.5 (8.8-20) | 0.055 |
| **Patte grade (%)**<br>1<br>2<br>3 | 0 | 34.5<br>48.3<br>17.2 | 44.8<br>34.5<br>20.7 | 39.7<br>41.4<br>18.9 | 0.563 |
| **Work as cause of injury (%)** | 0 | 96.6 | 17.2 | 56.9 | **<0.001** |
| **On leave at time of diagnosis (%)** | 1 | 10.7 | 34.5 | 22.8 | 0.07 |
| **Concomitant shoulder injury (%)** | 0 | 62.1 | 20.7 | 41.4 | **0.002** |
| **Duration of symptoms (weeks) (median, IQR)** | 0 | 59.1 (16.6-135.6) | 16.7 (7.1-56.6) | 28.3 (11.8-102.5) | **0.02** |
| **Undergone previous treatment (%)** | 0 | 31 | 44.8 | 37.9 | 0.42 |
| **Single-row Repair (%)**<br>Single | 0 | 37.9 | 51.7 | 44.8 | 0.34 |
| **Adjunct procedures (%)**<br>Biceps tenodesis | 0 | 51.7 | 75.9 | 63.8 | 0.07 |
| **Presentation to surgery (weeks) (median, IQR)** | 0 | 17 (7.1-43.6) | 4.7 (1.9 - 10.6 ) | 9.5 (4.2-30.7) | **<0.001** |

CP = compensable patients; non-CP = non-compensable patients

### Return to work outcomes

83% of patients returned to work in some capacity following RC repair. Significant differences in RTW status was observed (Table 3), with non-CP returning to pre-injury duties at a higher rate and a higher proportion of CP failing to return (Table 3). Of those that did RTW, the average time was 4.7 weeks (IQR 2.4-8.6) and was highly variable (Figure 2A), with no significant differences observed between groups (Table 3). A stepwise regression model explained 20.1% of the variance (adjusted) in first RTW time in patients that did return (N=49), with post-injury activity level (F=9.0, df=1, P=0.004), presence of comorbidities (F=2.4, df=1, P=0.13) and smoking status (F=8.1, df=1, P=0.007) included in the final model (Figure 2). Importantly, compensation status (Group) was not a significant factor.

**Table 3:** First RTW status and period from surgery to first RTW (separated by compensation status), and the final regression model for surgery to first RTW duration following RC repair.

| Outcomes | CP | Non-CP | All | P-value |
| --- | --- | --- | --- | --- |
| **First RTW status (%)** | N=29 | N=29 | N=58 | **<0.001** |
| Did not return | 17.2* | 0 | 8.6 |  |
| Pre-injury duties | 3.4 | 51.7* | 27.6 |  |
| Partial duties | 79.3 | 31.1 | 55.2 |  |
| Not working | 0 | 17.2 | 8.6 |  |
| **Surgery to first RTW time (weeks) (median, IQR)** | N=25<br>5.1 (2.6 - 8.6) | N=24<br>4.4 (0.7 - 11.1) | N=49<br>4.7 (2.4 - 8.6) | 0.86 |

\*largest contribution to $\chi^2$ ; CP = compensable patients; non-CP = non-compensable patients

**Figure 2:**
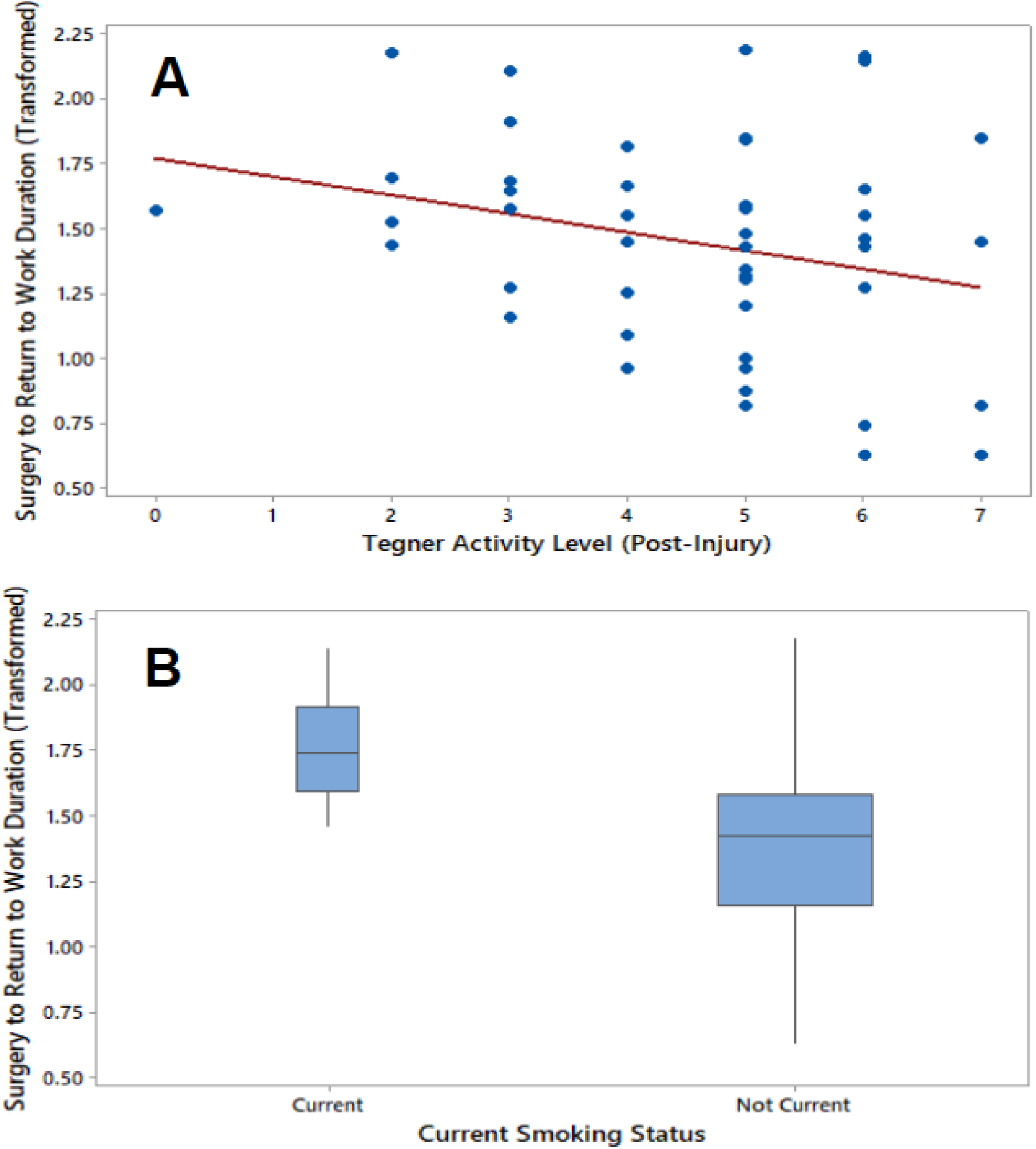
(A) Relationship between time to first RTW* and post-injury activity level. (B) Effect of smoking status on time to first RTW *Transformed with a Box-Cox approach

## Discussion

This study aimed to determine factors associated with first RTW status and time in patients undergoing aRCR under and outside an Australian workers compensation scheme, as the effects of modifiable factors on RTW in this population remain unexplored. This is the first study comparing outcomes of aRCR in a compensable patient group, compared to a set of age- and gender-matched controls. Of the CP cohort (N=29), 83% returned to work in some capacity, but only 3% first returned to pre-injury duties at an average of 5.1 weeks. Larger proportions of CP cohorts (64-94%) have been reported to return to pre-injury duties or normal activities by 7.6-9.8 months [9–11], however, it appears that return to pre-injury duties was examined as the final, not first RTW status in these studies.

In Australia, median time away from work for serious compensation claims in 2015-16 was 5.8 weeks [2], longer than the study median of 4.7 (N=49). The only systematic review and meta-analysis published investigating RTW in shoulder arthroplasty internationally (N=447) found a 64% rate of RTW by 2.3 months [20]. RTW rates reported in literature vary: 7% with twice as much time needed [21], 42% versus 94% in the standard population [22], 68% in a manual labour population [23], 82% with a change in work position often needed [24] and 90% for a younger population operated by arthroscopy [25,26]. A key reason for discrepancies is the variation in definitions of RTW status, which presents a considerable challenge when comparing RTW as an outcome.

In this investigation, first RTW status in any capacity was assessed due to the retrospective study design and lack of longer-term follow-up data. While RTW status was different between groups, time taken to first RTW did not differ between them. When considering the effect of compensation tiers on time away from work [12], significantly longer time away from work has been reported for highly compensated patients, indicating some effect of compensation status on time away from work, but was not evident in this cohort.

Smoking was an important factor which determined time taken to RTW, with current smokers taking longer to first to RTW compared to non-current smokers (Figure 2B). Longer symptom duration was also significantly associated with longer time of first RTW. Preoperative alcohol consumption, female gender, postoperative bursitis and heavy manual labour have been associated with poorer outcomes and increased time away from work [9–12], but these effects were not observed in this study.

Elucidating modifiable factors affecting RTW in the context of a compensation scheme requires further work. There has been limited examination of factors specific to RTW in aRCR, and current literature lacks adequately powered case-control studies with prospectively collected outcome-centred data. The use of outcome variables also requires some level of standardisation, pertaining to first and final RTW status and the definition of RTW capacity. A deeper understanding of the compensation process, and its interaction with modifiable factors affecting RTW outcomes is also required. Data generated from compensation claim forms is currently inadequate to predict failure to RTW within the NSW workers compensation scheme [15]. Key targets for intervention identified in this investigation are smoking status, and the preoperative activity level of the patient. However, further work is required to establish whether interventions designed to modify these factors are able to favourably affect patient outcomes.

This study is, to the best of the authors knowledge, the first to report a case-control series, and the first to review an Australian cohort receiving aRCR under a compensable scheme. The novelty of the results however, must be interpreted in light of certain limitations. Firstly, the observational design, combining retrospective chart review with phone follow-up is a weaker design in the present context. However, care was taken to match between CP and non-CP patients for known characteristics (age and gender). Nevertheless, future work should incorporate more robust prospective designs to establish stronger quality evidence for findings described here. Secondly the low consent rate (∼30%) from the CP group dictated a relatively small sample size compared to the published data to-date. Significant associations between outcomes and patient and postoperative management factors have been found that align with published literature, however care has been taken to not overfit the regression model on a small sample size.

There remains unexplained variance in the results, which may, to some degree, be accounted for by reporting, recruitment and performer bias. The patient groups were documented differently with work cover certificates used for CP patients and postoperative surgical correspondence used for non-CP patients (reporting bias), CP patients may have been influenced by the belief that participation may impact their claim status or established economic benefits (recruitment bias), and CP patients had better access to postoperative rehabilitation including physiotherapy and exercise physiology (performer boas). The effect of changes to the surgical procedure over a decade, though seemingly irrelevant, also cannot be disregarded. Future work requires clearer definitions of outcomes variables and timepoints, and would be greatly improved with a prospective study design with higher quality patient-centred data.

In conclusion, patients receiving aRCR under workers compensation schemes in one Australian jurisdiction are at increased risk of not returning to work compared to patients presenting outside a compensable scheme or in equivalent schemes internationally. Regardless of compensation status however, smokers and those with lower preoperative activity levels are at an increased risk of extended time to first return to work after aRCR. Further effort is required to determine if interventions aimed at altering the modifiable risk factors may improve outcomes for these patients.

## Supporting information

Supplementary material: postoperative protocol

STROBE checklist

## Data Availability

Data may be made available from the authors on reasonable request.

## Author contributions

IK, CS and JH contributed to the conceptualisation and stewardship of the study; MF and CS contributed to the study planning; VK conducted patient recruitment and data collection; JH supervised data collection; CS conducted the statistical analysis; MF drafted the manuscript, with assistance from CS and K.M.P; IK, VK and JH provided clinical feedback to the manuscript draft; IK sponsored the study.

## Acknowledgements

The authors would like to acknowledge Sofie French, Mac Cowley and Jessica Boh for their assistance with study planning and administration; Milad Ebrahimi for data collection coordination and his assistance with statistical analysis; Emily Zhong for her assistance with data collection; Ganesh Navaneedhan and Dinesh Choudary for their assistance with patient recruitment and data collection; and Meredith Harrison-Brown for their assistance with manuscript preparation.

