## Supplementary material: postoperative protocol for "Factors affecting first return to work following arthroscopic rotator cuff repair: A retrospective observational study in an occupational medicine clinic and single-surgeon practice"

### Post-Op Care Instructions and Exercises for Shoulder Arthroscopy

Patient Name: ______________________________ Date: ______________

Findings

______________________________________________
______________________________________________
______________________________________________
______________________________________________

Operation Performed

______________________________________________
______________________________________________
______________________________________________
______________________________________________

#### POST OPERATIVE CARE

Sling: Your affected arm will be placed in a fi tted sling or shoulder immobiliser. The shoulder immobiliser is designed to take the weight of your arm and keep your elbow away from your body. This will help the repair heal and decrease the pain so it is important that you relax your muscles when in it. The immobiliser may be taken off when showering, sitting or lying down, but the arm must be supported away from your body by your carer or by pillows. You will need to wear your immobiliser or sling for _______ weeks as instructed by Dr Herald.

At Home: You can use your arm in the sling for light activities such as writing, typing and using a knife and fork. It is important to come out of the sling 4-5 times per day to straighten your elbow and exercise your wrist and fi ngers to keep your joints mobile and strong. Squeezing a ball is a good way to do this.

Dressings: Keep the outer bandage on for 1 to 2 days if it stays dry and clean. After this you can remove it. To keep the outer bandage dry please shower with a plastic bag. Once removed the underneath dressings are waterproof but if soiled see me or your GP to get them changed as soon as possible.

Sutures: You will have buried, dissolving skin sutures that do not need to be removed.

Analgesia: A prescription has been provided by either myself, my assistant or my anaesthetist for analgesics. Please be aware that codeine containing products such as Panadeine Forte may cause constipation and drowsiness and should be used sparingly and with a high fi bre diet e.g. Metamucil.

| | |
|---|---|
| Rehabilitation: | Post-operative physiotherapy and home exercises are very important after surgery. Please follow the instructions shown later. You will be given a referral to formal physiotherapy which you should begin as soon as you can after leaving hospital. Without daily exercises you may end up with a stiff elbow. Please take your surgical booklet with you to your physiotherapy appointment. |
| Follow-up: | You will also need to contact Orthopaedic Clinic Sydney on 02 9233 3946 to arrange a follow-up appointment to see me at around 10 to 14 days. If it is not possible to see me, please see your GP at this time. |
| Driving a car: | You are not allowed to drive a car home after today's procedure. You are not safe to drive a car while your arm is in a sling. |

When to Worry:

- If you think you have an infection,
- Abnormal bleeding, a wound problem,
- A bandage that is too tight and cutting off your circulation,
- New numbness and tingling or any other emergency please contact the rooms immediately or failing that go back to hospital or see your GP.

Additional Instructions:

I look forward to seeing you again at your next appointment.

Dr Jonathan Herald
Orthopaedic Surgeon

### After Your Arthroscopy

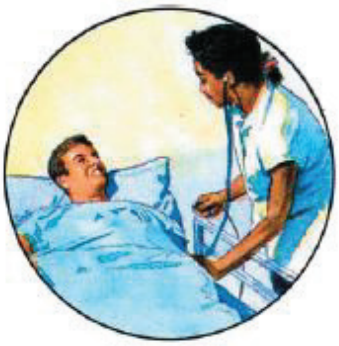

Arthroscopy is only part of the solution to a problem shoulder. You play a major role: rehabilitation is essential to speed your healing and get you back to normal activities. Immediately after surgery, you'll spend a few hours in a recovery area. You will be given a sling, an ice pack, and pain medications to make you more comfortable. Once home, follow the previous advice about staying comfortable and taking steps to speed your healing. You will be referred to physiotherapy soon after surgery to improve your shoulders range of motion and strength.

#### After Surgery

During the first few days after surgery, do what you can to stay comfortable. Ice your shoulder three times a day for about 20 minutes, and use a sling and pain medications. Move your arm and shoulder gently to help prevent stiffness and swelling. You will have a dressing change during your follow-up visit with me. If approved you may return to light work within a few days.

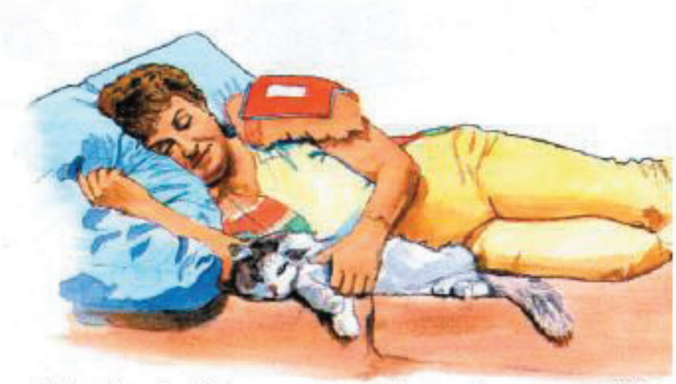

**A dry towel** will keep your wounds dry when you apply ice.

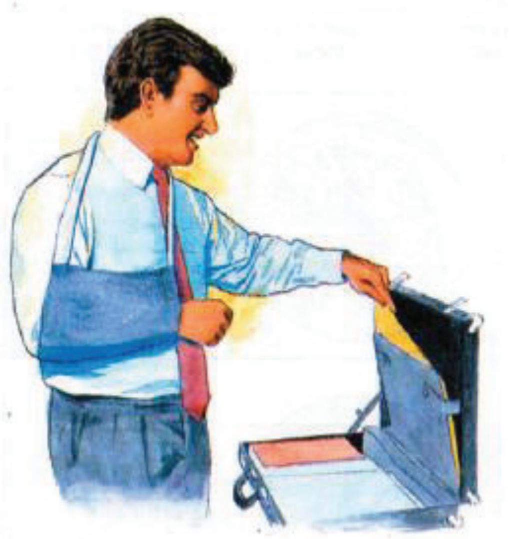

**The sling** will help reduce pain by resting the joint.

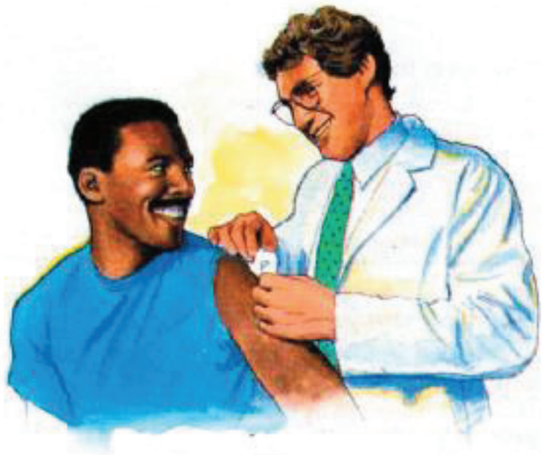

**Your dressings** will be changed during a follow-up visit.

### After Your Arthroscopy

#### Physiotherapy

You will have a referral for a program of exercise and other therapy beginning as early as the first week after surgery. Designed to get you back to normal activities, your personalised program is tailored to your specific type of shoulder problems and surgery. Stretches will help restore your shoulder's range of motion. When it is comfortable for you, your physiotherapist will teach you strengthening exercises.

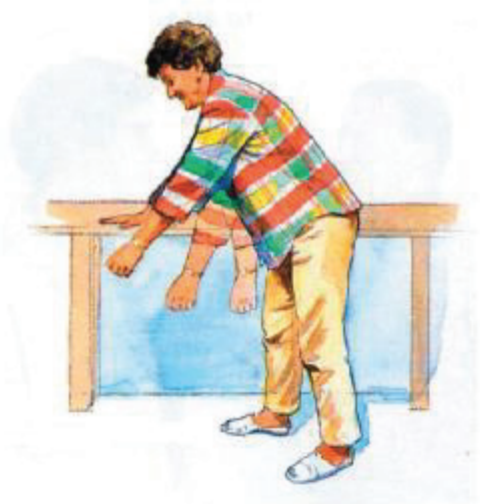

Restoring range of motion

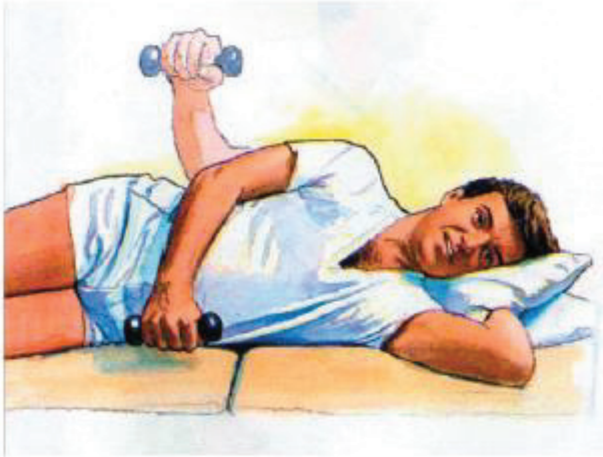

Increasing strength

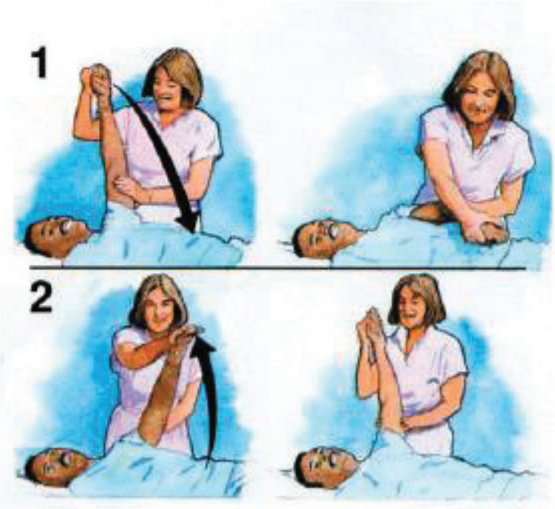


Increasing overhead strength and range of motion

### Personal Exercise Program

Shoulder Passive Range of Motion

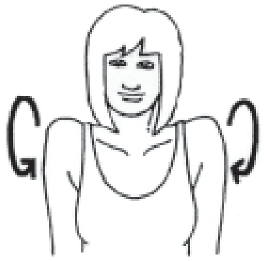

Sit or stand. Roll your shoulders in both directions. Repeat 10 times.

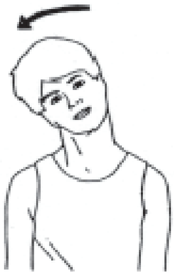

Sitting. Tilt your head toward one shoulder until you feel the stretch on the opposite side.
Hold approx. 5 seconds. Repeat on the other side.
Repeat 10 times.

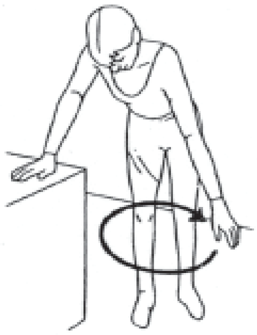

Stand leaning on a table with one hand.
Let your other arm hang relaxed straight down.
Swing your arm as if drawing a circle on the floor.
Change direction. Repeat 10 times.

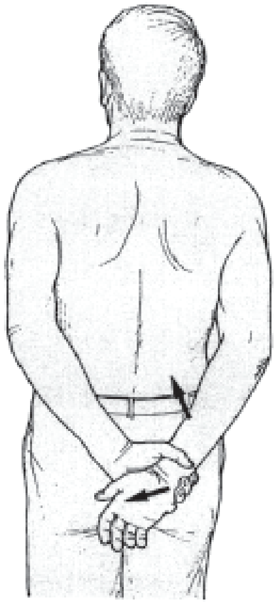

Stand. Hold onto the wrist of your other hand behind your back. Keep your elbow close to your body. Pull your wrist up and away from your back.
Repeat 10 times.

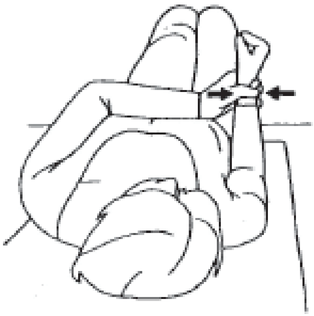

Lying on your back. Elbow bent at a right angle close to your body. Hold onto your wrist with your other hand. Move your arm outwards and stop when your forearm is perpendicular to your body then move your arm back inwards. Use the non-affected hand for movement. Repeat 10 times.

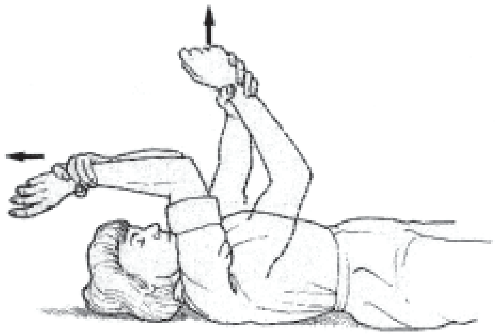

Lying on your back. Use one arm to lift the other arm up, keeping it as close to the ear as possible.
Repeat 10 times.
